# Routine implementation of α-synuclein Seed Amplification Assays reveals high diagnostic performance and the limited value of Alzheimer disease fluid biomarkers for detecting α-synuclein co-pathology

**DOI:** 10.64898/2026.04.21.26351389

**Authors:** Océane Jourdan, Marie Duchiron, Joan Torrent, Cédric Turpinat, Etienne Mondesert, Germain Busto, Mehdi Morchikh, Morgan Dornadic, Constance Delaby, Christophe Hirtz, Lou Thizy, Genevieve Barnier-Figue, Florence Perrein, Snejana Jurici, Audrey Gabelle, Karim Bennys, Sylvain Lehmann

## Abstract

**Background:** In neurodegenerative diseases, the diagnostic value of α-synuclein seed amplification assay (αSAA) in routine clinical practice and its relationship with cognitive performance and fluid biomarkers remains incompletely characterized. In addition, although Alzheimer disease (AD) CSF and blood biomarkers accurately detect amyloid and tau pathology, their ability to identify concomitant α-synuclein (αSyn) pathology remains unknown.

**Methods:** We included 398 patients from the multicenter memory clinic ALZAN cohort. All participants underwent CSF and blood sampling with measurement of CSF biomarkers (Aβ42/40, tau, p-tau181) and plasma biomarkers (Aβ42/40, p-tau181, p-tau217, GFAP, NfL). Cognitive assessment was performed using the Mini-Mental State Examination (MMSE). Clinical diagnoses were independently confirmed by two senior neurologists. αSyn status was determined by αSAA (RT-QuIC).

**Results:** Of 398 patients, 19/20 patients with Lewy body dementia (LBD) (95.0%) and 32/203 patients with AD (15.8%) were αSAA+. αSAA-positivity presented a sensitivity of 95.0% (95%CI, 75.1-99.9), a specificity of 93.1% (95%CI, 88.3-96.4) for distinguishing LBD from patients without LBD or AD. In the entire cohort, αSAA+ patients showed lower MMSE scores (p<0.01), lower CSF Aβ42/40 ratio (p<0.01), and elevated plasma GFAP (p<0.05). However, within the AD subgroup, only CSF Aβ42/40 differed marginally between αSAA+ and αSAA- patients, whereas CSF total tau, CSF p-tau181, plasma p-tau217, p-tau181, GFAP, and NfL showed no significant differences.

**Conclusions:** αSAA accurately detects αSyn pathology in routine memory clinic practice that is not captured by current AD CSF and blood biomarkers. These findings support αSAA as a complementary biomarker for routine diagnosis and patient stratification.

**What is already known on this topic:** α-synuclein seed amplification assay (αSAA) has shown high sensitivity and specificity for detecting Lewy body dementia (LBD) in research cohorts, and α-synuclein co-pathology is frequently observed in Alzheimer’s disease (AD). However, it remains unknown whether currently available AD CSF and blood biomarkers can identify patients with α-synuclein pathology in routine clinical practice.

**What this study adds:** In a prospective multicenter memory clinic cohort, αSAA showed high sensitivity (95.0%) and specificity (93.5%) for identifying clinically diagnosed LBD and detected αSyn co-pathology in 15.8% of AD patients. Current AD CSF and blood biomarkers did not discriminate between αSAA-positive and αSAA-negative patients, providing little information on concomitant αSyn pathology.

**How this study might affect research, practice or policy:** αSAA provides unique biological information by improving the detection of LBD and identifying AD patients with concomitant αSyn co-pathology that is not captured by current CSF and blood biomarkers, thereby improving patient stratification and informing future precision medicine approaches and therapeutic trials.

## Introduction

A key characteristic of neurodegenerative diseases is the accumulation of misfolded proteins with altered physicochemical properties, such as amyloid β-peptides (Aβ) and tau proteins in Alzheimer disease (AD), collectively referred to as proteinopathies ^1^. Synucleinopathies, including Lewy body dementia (LBD), are similarly neuropathologically defined by the presence of α-synuclein (αSyn) aggregates in the brain, a major component of Lewy bodies and Lewy neurites ^2^ ^3^.

An important challenge in neurodegenerative disease diagnosis is the frequent co-existence of multiple pathological proteins within the same brain ^4^ ^5^. For example, αSyn deposits are observed in 20-50% of patients with neuropathologically confirmed AD ^6-8^, while 28-89% of LBD patients exhibit tau deposits and Aβ plaques ^9^. This co-pathology, common in older individuals, may result from interactions between misfolded proteins and could contribute to the severity and heterogeneity of clinical manifestations observed in both AD and LBD patients ^8^. Importantly, despite major advances in AD fluid biomarkers, currently available CSF and blood biomarkers accurately detect amyloid and tau pathology but provide little information on concomitant αSyn pathology.

The challenge of detecting αSyn pathology has limited our ability to characterize this co-pathology in living patients. Consequently, synucleinopathies are mainly suspected on clinical features and imaging markers. Gold standard diagnosis requires identification of αSyn aggregates in *postmortem* brain tissue ^10^. Recent advances have demonstrated that misfolded αSyn can be detected in cerebrospinal fluid (CSF) and other tissues using *in vitro* seed amplification assays (SAAs) ^11^. Originally developed as part of research on prion diseases, these techniques enable the qualitative detection of αSyn *in vitro* self-propagated aggregation. αSyn SAAs (αSAA), particularly real-time quaking-induced conversion (RT-QuIC), have shown high diagnostic accuracy in detecting αSyn pathology and differentiating synucleinopathies from other neurodegenerative diseases, as confirmed by *postmortem* neuropathological studies ^12-16^.

While blood and CSF biomarkers have transformed the diagnosis of AD, it remains poorly investigated whether they also capture concomitant αSyn pathology. However, implementing αSAA testing in routine clinical practice could improve diagnostic accuracy and enable detection of αSyn co-pathology in AD patients. Nevertheless, the clinical utility of αSAA in routine memory clinic settings and its added value relative to established AD fluid biomarkers is still incompletely characterized.

Whether current AD fluid biomarkers can identify patients with αSyn co-pathology remains largely unknown. Addressing this question is important with regards to the implementation of αSAA as a complementary biomarker in routine diagnostic.

In the present study, we used the prospective multicenter ALZAN memory clinic cohort to evaluate the diagnostic performance of αSAA in routine clinical practice and to determine whether currently available AD CSF and plasma biomarkers can identify αSyn pathology. Specifically, we (1) assessed the diagnostic performance of αSAA for distinguishing clinically diagnosed LBD from other dementias; (2) determined the prevalence of αSyn co-pathology in AD patients; and (3) examined the associations between αSAA positivity and established CSF and plasma biomarkers of amyloid, tau, and neurodegeneration.

## Methods

### Study population and selection criteria

The multicenter prospective ALZAN cohort (ClinicalTrials.gov Identifier #NCT05427448) included adult patients recruited from memory clinics at the hospitals of Montpellier, Nimes, and Perpignan between November 2022 and July 2024. A total of 398 patients were enrolled, each providing written informed consent for the collection and use of their biological samples for research purposes. Eligible participants were adults presenting with cognitive impairment undergoing diagnostic evaluation in a memory clinic who provided written informed consent and underwent concomitant CSF and blood sampling. Exclusion criteria included uncontrolled medical conditions that could interfere with study participation, recent participation in other clinical trials that could confound results, and refusal to provide consent. Clinical and demographic data collected at baseline included age, sex, body mass index (BMI), and Mini-Mental State Examination (MMSE) score. The MMSE was performed within routine clinical assessment according to national guidelines ^17^.

Clinical diagnoses were established according to the following guidelines: the National Institute on Aging–Alzheimer’s Association criteria for AD McKhann ^18^, the International Consortium criteria for frontotemporal dementia (FTD) ^19^, and the Fourth LBD Consortium criteria for LBD ^10^. Participants were categorized into four clinical diagnostic groups: AD, LBD, FTD, and Other (Figure 1). The Other diagnostic group was heterogeneous, encompassing atypical tauopathies, vascular dementia, psychiatric disorders, subjective and mild cognitive impairment, inflammatory/infectious conditions, and other non-neurodegenerative causes (see Supplementary Table 1 for the complete diagnostic breakdown). Participants with missing diagnoses were excluded from these clinical diagnostic groups.

**Figure 1.**
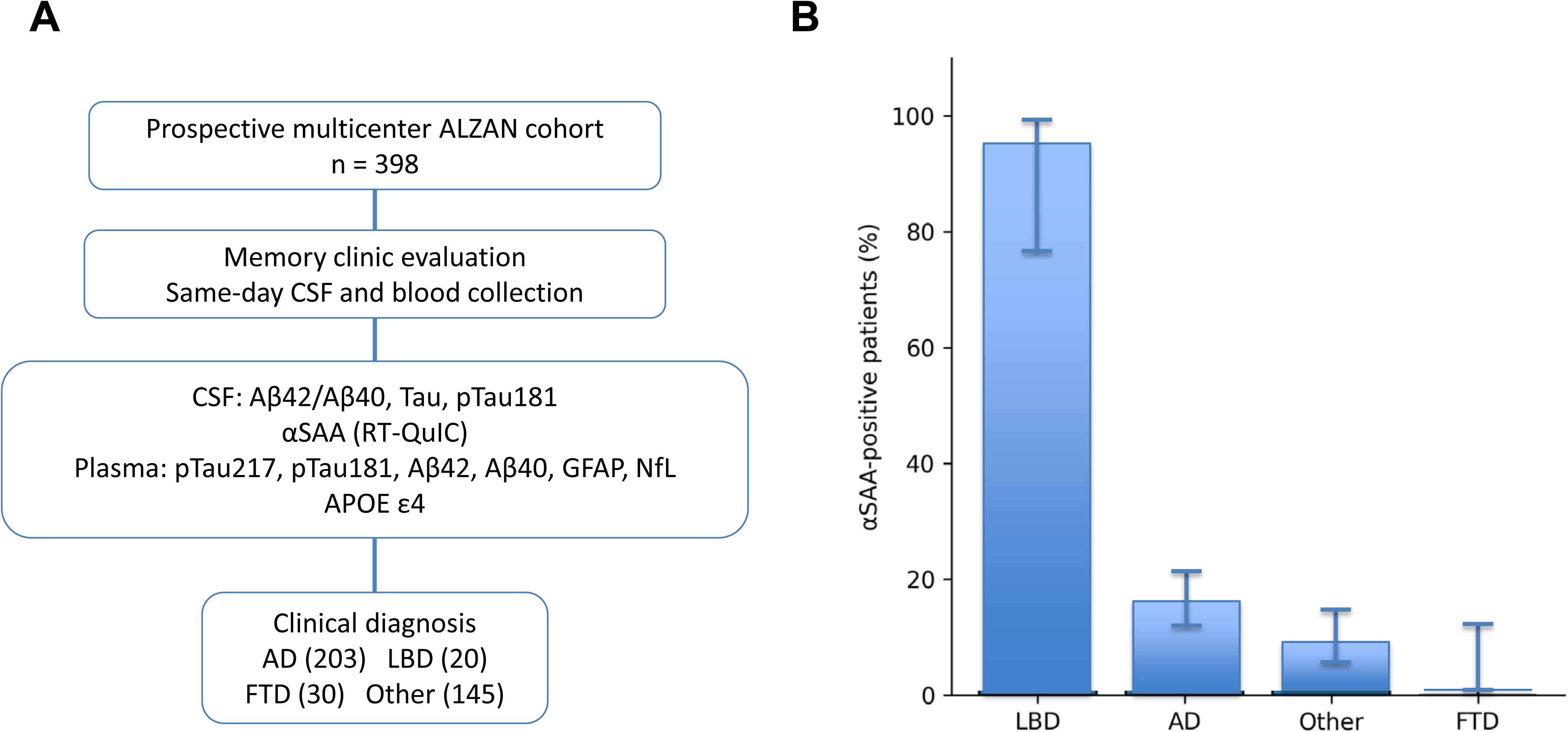
Study design and distribution of αSAA positivity **Panel A :** Patients referred to three French memory clinics underwent standardized clinical assessment, same-day cerebrospinal fluid (CSF) and blood sampling, and biomarker analyses including αSAA by real-time quaking-induced conversion (RT-QuIC). Clinical diagnoses were established according to international consensus criteria independently of αSAA results. **Panel B :** Proportion of αSAA-positive patients according to the final clinical diagnosis. Error bars represent 95% confidence intervals. **Abbreviations:** AD, Alzheimer disease; LBD, Lewy body dementia; αSAA, α-synuclein seed amplification assay.

### CSF collection and analysis

CSF collection, processing, and storage were performed according to a standardized procedure in line with international consensus guidelines ^20^. CSF concentrations of Aβ (1-40) and (1-42) (Aβ40 and Aβ42), total tau (tau), and phosphorylated-tau181 (p-tau181) were measured using Lumipulse G1200® system (Fujirebio France). The CSF Aβ42/Aβ40 ratio was used as a biomarker of cerebral amyloid pathology, with a cut-off set at 7% ^21^.

### Blood biomarkers

Blood samples were collected on the same day as the lumbar puncture and processed at 2,000 g for 10[minutes. Plasma aliquots were stored at -80 °C until analysis. The estimated glomerular filtration rate (eGFR) was calculated using the 2021 CKD Epidemiology Collaboration (CKD-EPI) equation^22^. Phosphorylated-tau217 (p-tau217) was measured using commercial kits on Lumipulse G1200® system (Fujirebio France) ^23^. P-tau181, Aβ42, Aβ40, glial fibrillary acidic protein (GFAP), neurofilament light chain (NfL), and apolipoprotein E ε4 allele (APOE ε4) carrier status were measured using Elecsys® assays on the Cobas e402 automated system (Roche Diagnostics) ^24^.

### Synuclein seed amplification assay (αSAA) protocol

For αSAA analysis, we used a real-time quaking-induced conversion (RT-QuIC) approach, adapted from the method described by Verdurand et al. ^25^, with all analyses performed in a single laboratory (Montpellier, France). Recombinant human αSyn was produced locally and purified according to the protocol described by Pujols et al. ^26^.

RT-QuIC assays were performed in 96-well plates and monitored in real time using a FLUOstar Omega plate reader (BMG Labtech). Plates were incubated at 42°C under intermittent shaking cycles (400 rpm; 1 min on/1 min off). Thioflavin T fluorescence was measured every 45 minutes (excitation: 448 nm; emission: 482 nm) over a period of 168-225 hours. Each CSF sample was analyzed in quadruplicate with positive and negative controls. A sample was considered αSAA positive (αSAA+) when at least one of the four replicates exceeded a predefined fluorescence threshold (corresponding to 50% of the mean difference between the aggregation and baseline fluorescence signals of the positive control). Samples with no positive wells were classified as SAA-negative (αSAA–).

From the fluorescence kinetics, three quantitative parameters were extracted: the lag phase (Lag), defined as the time required for fluorescence to reach twice the baseline value, indicating seeding, the maximum fluorescence intensity (FluoMax) and the area under the fluorescence curve (Area). The number of positive replicates per sample was also recorded. The αSAA assay was analytically validated according to applicable quality standards before clinical implementation. Validation results are summarized in Supplementary Figure 1 and Supplementary Table 2. Clinical diagnoses were established independently of αSAA results, and laboratory personnel remained blinded to all clinical information throughout the analyses.

### Statistical analysis

Population characteristics and αSAA quantitative parameters across clinical diagnostic groups were described using percentages (%) for categorical variables and means with standard deviations (mean ± SD) for continuous variables. Comparisons were performed using the χ² test for categorical variables, Student’s t-test for comparisons between two groups, and one-way analysis of variance (ANOVA) followed by Tukey’s honestly significant difference (HSD) post hoc test for multiple-group comparisons. All statistical tests were two-sided, and statistical significance was set at p < 0.05. Receiver operating characteristic (ROC) curves were constructed using clinical LBD diagnosis as the reference standard. Areas under the curve (AUCs) were reported with 95% confidence intervals (CIs). All analyses were performed using MedCalc (ver. 20.118) and R software (ver. 4.4.2).

## Results

### Characteristics of the ALZAN population

In the ALZAN cohort of 398 participants, 203 (51%) were classified as AD, 20 (5%) as LBD, 30 (7.5%) as FTD, and 145 (34.7%) as Other. The mean age at inclusion was 71.1 ± 10.5 years. Age differed significantly across groups (p < 0.0001), with younger patients in the FTD group (68.3 years) and older patients in the LBD group (76.6 years). Women comprised 52.5% (n = 209) of the cohort, with a higher proportion in AD (60.1%) and a lower proportion in LBD (30.0%). MMSE scores differed significantly across groups (p < 0.0001), with lower scores in AD than in the Other group (23 vs 26, respectively; p < 0.0001). Full cohort characteristics are presented in Table 1.

**Table 1.** Characteristics of the overall cohort and by clinical diagnostic group: demographics, cognition, and biomarkers (n = 398). Data are presented as mean (M ± SD) for continuous variables and n (%) for categorical variables. One-way ANOVA followed by Tukey’s HSD post-hoc analysis was used for continuous variables; χ² test for categorical variables. Lag (hours), Fluomax, Area, and Npositive refer to αSAA parameters. P-values are indicated in the last column (*p < 0.05; **p < 0.01; ***p < 0.001).

| Criteria | Whole population<br>(n=398) | AD (n=203) | LBD (n=20) | FTD (n=30) | Other<br>(n=145) | p-value |
| --- | --- | --- | --- | --- | --- | --- |
| Age, years, M (SD) | 71.1 (10.5) | 73.7 (7.6) | 76.6 (6.3) | 68.3 (8.6) | 67.5 (12.8) | p<0.0001*** |
| Sex, female, n (%) | 209 (52.5) | 122 (60.1) | 6 (30.0) | 18 (60.0) | 61 (44.2) | p=0.0042 ** |
| BMI (kg/m<sup>2</sup>), M (SD) | 24.7 (4.1) | 24.2 (3.9) | 24.5 (4.3) | 24.6 (3.7) | 25.6 (4.3) | p=0.0311 * |
| MMSE (/30), M (SD) | 23.1 (4.6) | 21.9 (4.7) | 22.3 (4.9) | 23.2 (4.4) | 25.4 (3.6) | p<0.0001*** |
| eGFR (mL/min/1.73m<sup>2</sup>), M (SD) | 84.7 (15.8) | 84.6 (14.6) | 78.4 (13.4) | 81.9 (14.2) | 86.1 (17.4) | p=0.1609 |
| CSF Aβ40 (pg/mL), M (SD) | 10856 (3896) | 12214 (3568) | 9926 (3450) | 10795 (4187) | 9210 (3621) | p<0.0001*** |
| CSF Aβ42 (pg/mL), M (SD) | 733.1 (356.8) | 600.2 (192.9) | 702.4 (242.3) | 1019.2 (454.5) | 877.1 (437.3) | p<0.0001*** |
| CSF Aβ42/40 (%), M (SD) | 7.1 (2.8) | 5.1 (1.5) | 7.5 (2.6) | 9.4 (1.9) | 9.4 (2.2) | p<0.0001*** |
| CSF tau (pg/mL), M (SD) | 488.8 (352.8) | 675.6 (346.8) | 345.3 (182.6) | 354.5 (215.0) | 280.5 (246.8) | p<0.0001*** |
| CSF p-tau181 (pg/mL), M (SD) | 71.2 (58.1) | 106.7 (60.7) | 50.8 (31.7) | 41.1 (24.9) | 30.9 (13.6) | p<0.0001*** |
| Pl GFAP (pg/mL), M (SD) | 135.2 (63.7) | 155.3 (58.1) | 142.4 (51.4) | 116.9 (63.4) | 108.9 (61.2) | p<0.0001*** |
| Pl p-tau217 (pg/mL), M (SD) | 0.41 (0.42) | 0.62 (0.44) | 0.20 (0.10) | 0.18 (0.18) | 0.16 (0.19) | p<0.0001*** |
| Pl Aβ40 (pg/mL), M (SD) | 296.8 (52.7) | 299.9 (46.7) | 294.5 (55.1) | 288.0 (49.5) | 293.8 (58.6) | p=0.5505 |
| Pl Aβ42 (pg/mL), M (SD) | 35.0 (8.4) | 32.3 (6.7) | 38.0 (7.0) | 38.1 (6.3) | 37.8 (9.4) | p<0.0001*** |
| Pl Aβ42/40 (%), M (SD) | 11.9 (2.8) | 10.8 (1.9) | 13.9 (7.2) | 13.4 (1.8) | 12.9 (2.2) | p<0.0001*** |
| Pl p-tau181 (pg/mL), M (SD) | 1.3 (0.75) | 1.6 (0.71) | 0.96 (0.29) | 0.89 (0.39) | 0.90 (0.47) | p<0.0001*** |
| Pl NfL (pg/mL), M (SD) | 4.4 (4.9) | 3.9 (1.8) | 3.7 (1.7) | 7.4 (5.5) | 4.4 (7.3) | P=0.0031** |
| APOE ε4 carrier, n (%) | 171 (43.0) | 128 (63.1) | 6 (30.0) | 4 (13.3) | 32 (23.2) | p<0.0001*** |
| αSAA +, n (%) | 63 (15.8) | 32 (15.8) | 19 (95.0) | 0 (0) | 12 (8.3) | p<0.0001*** |

Abbreviations: AD, Alzheimer's disease; LBD, Lewy body dementia; FTD, frontotemporal dementia; BMI, body mass index; MMSE, Mini-Mental State Examination; eGFR, estimated glomerular filtration rate; CSF, cerebrospinal fluid; Pl, plasma; p-tau, phosphorylated tau; NfL, neurofilament light chain; αSAA, α-synuclein seed amplification assay

AD patients exhibited the expected biochemical profile, characterized by higher CSF tau and p-tau181 concentrations and lower Aβ42/Aβ40 ratio than LBD, FTD, and Other groups (all p < 0.0001). The proportion of APOE ε4 carriers was highest in the AD group (63.1%) compared with LBD (30.0%), FTD (13.3%), and Other groups (23.2%). Plasma NfL levels were higher in the FTD group than in AD, LBD, and Other groups (all p < 0.05).

Consistent with previous findings from the ALZAN cohort ^27^, plasma p-tau217 showed the highest diagnostic performance for distinguishing clinically diagnosed AD from non-AD participants (AUC, 0.902; 95% CI, 0.871–0.933), followed by the plasma Aβ42/Aβ40 ratio (AUC, 0.803; 95% CI, 0.758–0.848) (Supplementary Figure 2).

### Performance of αSAA and application to the ALZAN cohort

Within the ALZAN cohort, 63 of 398 patients (15.8%) were αSAA-positive. Positivity was markedly higher in the LBD group (19/20, 95.0%) than in the AD (32/203, 15.8%), Other (12/145, 8.3%), and FTD (0/30, 0%) groups (overall p < 0.0001) (Figure 1). Because αSyn co-pathology is common in AD, AD patients were excluded from the assessment of αSAA diagnostic performance for LBD. In the remaining cohort, αSAA identified clinically diagnosed LBD with a sensitivity of 95.0% (95% CI, 75.1–99.9), a specificity of 93.1% (95% CI, 88.3–96.4), a positive predictive value of 61.3% (95% CI, 47.6–73.4), and a negative predictive value of 99.4% (95% CI, 96.0–99.9).

### Biomarker levels associated with αSAA status

As an initial exploratory analysis, we compared all αSAA-positive (n = 63) and αSAA-negative (n = 335) participants irrespective of clinical diagnosis. αSAA-positive patients were older (73.9 vs 70.0 years, p=0.0013), had lower MMSE scores (p=0.0085), lower CSF Aβ42/Aβ40 ratio (p=0.0051), and higher plasma GFAP concentrations (p=0.0395) than αSAA-negative patients (Supplementary Table 3). However, because αSAA positivity was strongly enriched in patients with LBD, these differences were likely driven by the underlying diagnostic composition rather than by α-synuclein pathology itself.

To determine whether currently available Alzheimer disease fluid biomarkers identify αSyn co-pathology, we subsequently restricted the analysis to patients with clinically diagnosed AD. As shown in Table 2 and Figure 2, only the CSF Aβ42/Aβ40 ratio was lower in αSAA-positive AD patients, whereas CSF total Tau, CSF p-tau181, plasma p-tau217, plasma p-tau181, plasma Aβ42/Aβ40 ratio, plasma GFAP, and plasma NfL did not differ significantly according to αSAA status, indicating that current AD CSF and plasma biomarkers provide little information on concomitant α-synuclein pathology.

**Figure 2.**
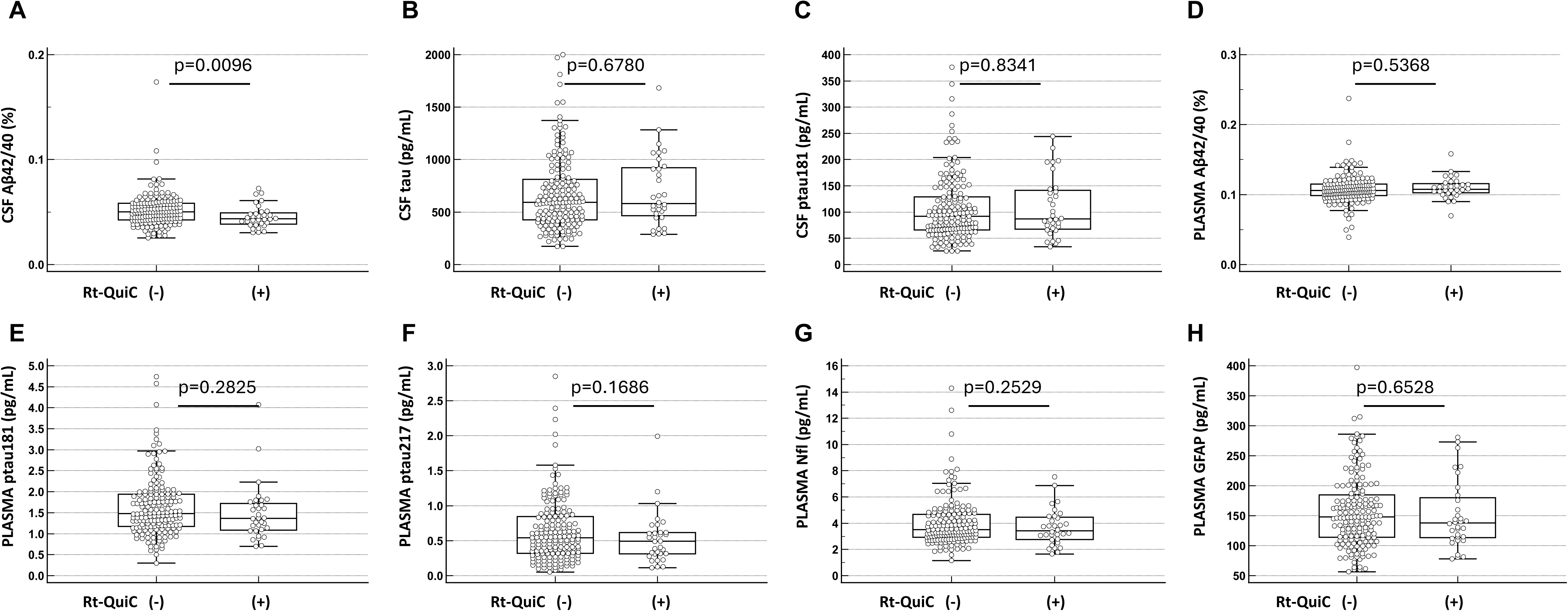
Biomarker profiles according to αSAA positivity on AD patients. Boxplots show the distribution of CSF Aβ42/Aβ40 ratio (A), CSF total Tau (B), CSF p-tau181 (C), plasma p-tau181 (D), plasma p-tau217 (E), plasma NfL (F), and plasma GFAP concentrations. Boxes represent the median and interquartile range (IQR). Comparisons between groups were performed using the Wilcoxon rank-sum test, with corresponding *p*-values displayed above each panel. No significant differences were observed between αSAA- and αSAA+ Alzheimer’s disease patients for any of the measured biomarkers, indicating that conventional CSF and plasma biomarkers of amyloid pathology, tau pathology, neurodegeneration, and astroglial activation do not distinguish AD patients with concomitant α-synuclein pathology detected by αSAA.

**Table 2.** Demographic, cognitive and biomarker results between AD+αSAA- and AD+αSAA+ patients (n = 198). Data are presented as mean (M ± SD) for continuous variables and n (%) for categorical variables. Differences between AD+αSAA- and AD+αSAA+ subgroups were assessed using Student’s-t-test for continuous variables and χ² tests for categorical variables. P-values are shown in the last column (*p < 0.05).

| Criteria | AD+αSAA- (n=171) | AD+αSAA+ (n=32) | p-value |
| --- | --- | --- | --- |
| Age (years), M (SD) | 73.9 (7.6) | 72.5 (7.3) | p=0.3077 |
| Sexe. Female, n (%) | 104 (60.8) | 18 (56.3) | p=0.6281 |
| BMI (kg/m<sup>2</sup>), M (SD) | 24.3 (4.0) | 23.3 (3.5) | p=0.1464 |
| MMSE (/30), M (SD) | 22.1 (4.7) | 21.2 (3.9) | p=0.2707 |
| eGFR (mL/min/1.73m<sup>2</sup>), M (SD) | 84.3 (14.9) | 86.6 (13.3) | p=0.3758 |
| CSF Aβ40 (pg/mL), M (SD) | 12087 (3455) | 12892 (4116) | p=0.3047 |
| CSF Aβ42 (pg/mL), M (SD) | 606.7 (200.9) | 565.3 (139.8) | p=0.1607 |
| CSF Aβ42/40 (%),M (SD) | 5.2 (1.6) | 4.6 (1.1) | p=0.0096 ** |
| CSF tau (pg/mL), M (SD)) | 671.3 (349.9) | 698.4 (334.4) | p=0.6780 |
| CSF p-tau181 (pg/mL), M (SD) | 106.3 (61.4) | 108.7 (57.3) | p=0.8341 |
| Pl GFAP (pg/mL), M (SD) | 156.1 (58.5) | 151.1 (56.6) | p=0.6528 |
| Pl p-tau217 (pg/mL), M (SD) | 0.64 (0.45) | 0.53 (0.37) | p=0.1686 |
| Pl Aβ40 (pg/mL), M (SD) | 300.8 (48.4) | 295.4 (36.8) | p=0.4693 |
| Pl Aβ42 (pg/mL), M (SD) | 32.3 (6.9) | 32.3 (5.5) | p=0.9720 |
| Pl Aβ42/40 (%),M (SD) | 10.8 (2.0) | 11.0 (1.5) | p=0.5368 |
| Pl p-tau181 (pg/mL), M (SD) | 1.6 (0.71) | 1.5 (0.67) | p=0.2825 |
| Pl NfL (pg/mL), M (SD) | 4.0 (1.8) | 3.5 (2.9) | p=0.2529 |
| APOE ε4 carrier, n (%) | 106 (62.0) | 22 (68.8) | p=0.4670 |

Abbreviations: n, number of patients; αSAA, α-synuclein seed amplification assay; BMI, body mass index; MMSE, Mini-Mental State Examination; eGFR, estimated glomerular filtration rate; CSF, cerebrospinal fluid; Pl, plasma; p-tau, phosphorylated tau; NfL, neurofilament light chain; APOE ε4, apolipoprotein E ε4 allele; GFAP, glial fibrillary acidic protein.

### Biomarker profiles of αSAA-positive patients according to clinical diagnosis

To further characterize αSAA-positive patients (n = 63), we compared cognitive and biomarker profiles according to the underlying clinical diagnosis (AD, LBD, and Other; Table 3). As expected, CSF and plasma biomarkers of amyloid and tau pathology differed markedly across diagnostic groups, reflecting their underlying biological profiles. AD+αSAA+ patients exhibited the lowest CSF Aβ42/Aβ40 ratio (4.6% vs 7.3% in LBD and 9.2% in Other; p < 0.0001), the highest CSF tau (p < 0.0001) and p-tau181 (p < 0.0001) concentrations, the highest plasma p-tau217 concentration (p < 0.0001), and the highest frequency of APOE ε4 carriers (p=0.0032). Conversely, plasma NfL concentrations were highest in the Other+αSAA+ subgroup (p=0.0075). No significant differences were observed for age, MMSE, eGFR, plasma GFAP, plasma Aβ40, or plasma Aβ42/Aβ40 ratio. Overall, these findings indicate that the biomarker profile of αSAA-positive patients is primarily determined by the underlying clinical diagnosis rather than by αSyn positivity itself.

**Table 3.**
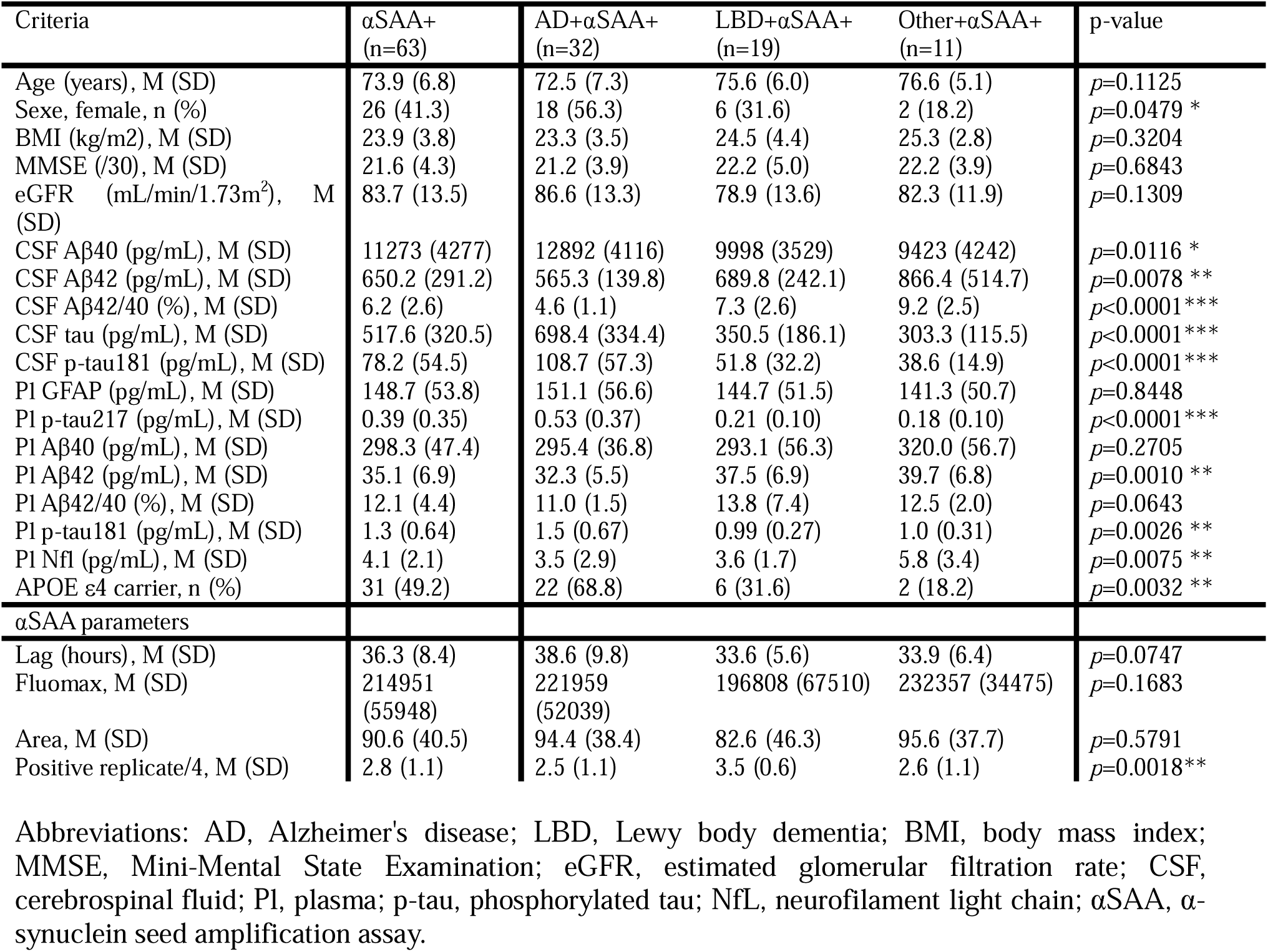
Demographic, cognitive, and biological biomarker results across αSAA+ patients according to clinical diagnosis (AD+αSAA+, LBD+αSAA+, Other+αSAA+; n=63). Data are presented as mean (M ± SD) for continuous variables and n (%) for categorical variables. Differences across diagnostic groups were assessed using one-way ANOVA with Tukey’s post-hoc analysis for continuous variables and χ² test for categorical variables. P-values are shown in the last column (*p < 0.05; **p < 0.01). The Other+αSAA+ subgroup (n=11) comprised patients with subjective cognitive impairment (n=3), atypical tauopathies (n=3), other neurodegenerative causes (n=3), inflammatory/infectious conditions (n=1), and atypical parkinsonian syndromes (n=1). Lag (hours), Fluomax, Area, and Npositive refer to αSAA parameters

Regarding αSAA kinetics, LBD patients exhibited a shorter lag phase than AD patients (p=0.0254). Across the three αSAA-positive diagnostic groups (Table 3), the number of positive replicates also differed significantly (p=0.0018), with higher values in LBD than in both AD (p=0.0017) and Other (p=0.0328) patients. In contrast, FluoMax and Area did not differ significantly between groups (p=0.1683 and p=0.5791, respectively).

### Characteristics and Effects of αSyn co-pathology in AD group

To determine whether current Alzheimer’s disease fluid biomarkers identify concomitant αSyn pathology, we compared αSAA-positive and αSAA-negative patients within the AD group. The two groups were comparable in age (*p=*0.3077), sex (*p=*0.6281), renal function (*p=*0.3758), APOE ε4 carrier status (*p=*0.4670), and cognitive performance as assessed by MMSE (*p=*0.2707), although MMSE scores tended to be lower in αSAA-positive patients (21.2 vs 22.1). A lower CSF Aβ42/Aβ40 ratio was initially observed in αSAA-positive AD patients (4.6% vs 5.2%; *p=*0.0096). However, no significant differences were found for CSF total Tau, CSF p-tau181, plasma p-tau217, plasma p-tau181, plasma Aβ42/Aβ40 ratio, plasma GFAP, or plasma NfL (Table 2 and Figure 2). Overall, these findings indicate that currently available AD CSF and plasma biomarkers do not distinguish AD patients with concomitant α-synuclein pathology detected by αSAA.

## Discussion

Our results show that 15.8% of patients in the ALZAN cohort tested positive for αSAA, with significant disparities across clinical diagnoses. The sensitivity for LBD detection was 95%, with a specificity of 93.5% and a negative predictive value of 99.4%. These results are consistent with previous clinical studies conducted in cognitive cohorts, reporting sensitivities ranging from 72.3% to 95.2% and specificities between 97% and 100% ^25^ ^28-31^. Overall, these results support the robust performance of αSAA for detecting αSyn pathology in LBD. Importantly, the sensitivity observed in our cohort is comparable to that reported in large, standardized, neuropathologically confirmed cohorts, with sensitivities of approximately 90% to 95% ^30-32^. Differences across studies seem likely influenced by cohort composition and diagnostic classification strategies, including limitations of clinical diagnostic frameworks such as McKeith criteria ^10^. Previous studies have highlighted variability in αSAA performance according to disease stage and the specific brain regions affected, which may influence the probability of aggregate amplification ^33^. In line with this, Tosun et al. ^31^ reported sensitivities ranging from 57% in limbic forms to 100% in neocortical forms. A major strength of our study is its reflection of real-world clinical practice in unselected patients, with pre-analytical variations ^34^ ^35^, demonstrating that αSAA can be reliably implemented in routine diagnostic workflows while maintaining excellent diagnostic performance.

In the ALZAN cohort, 15.8% of clinically diagnosed AD patients were αSAA-positive, consistent with previous in vivo studies reporting αSyn co-pathology in approximately 14-21% of AD patients, although lower than frequencies reported in neuropathological series ^8^ ^25^ ^28^. This finding highlights the clinical relevance of αSAA for identifying biologically defined AD subgroups that cannot be recognized on clinical grounds alone.

We also observed differences in the kinetic parameters of αSAA between AD and LBD patients, with a shorter lag phase and a higher number of positive replicates in LBD patients, in line with Verdurand et al. ^25^. This recently reported observation may reflect differences in the concentration and/or seeding efficiency of misfolded αSyn species in CSF across clinical phenotypes ^33^. However, αSAA kinetic parameters should not be interpreted as direct markers of specific types of αSyn pathology. Rather, lag phase, fluorescence, and related metrics likely reflect a combination of biological and assay-dependent factors rather than strictly disease-specific signatures ^36^ ^37^. Consequently, the substantial overlap between AD and LBD distributions limits their standalone discriminative performance.

A major finding of this study is that current AD CSF and plasma biomarkers failed to distinguish AD patients with concomitant αSyn pathology detected by αSAA. Although exploratory analyses across the whole cohort showed differences in MMSE, CSF Aβ42/Aβ40 ratio, and plasma GFAP, these differences were largely explained by the unequal distribution of clinical diagnoses. When analyses were restricted to patients with AD, no meaningful differences were observed for established biomarkers of amyloid pathology, tau pathology, neurodegeneration, or astroglial activation. These findings indicate that currently available AD fluid biomarkers provide little information on the presence of concomitant αSyn pathology.

Several studies indicate that AD patients with αSyn co-pathology exhibit a more severe clinical profile, including greater cognitive and motor impairments and faster decline ^28^ ^38^. At the biological level, available data remain limited and sometimes inconsistent. While Verdurand et al. ^25^ reported no significant differences between AD+αSAA− and AD+αSAA+ patients, Esteller-Gauxax et al. ^39^ observed differences in plasma p-tau217 and GFAP. Our results are overall more consistent with those of Verdurand et al., suggesting that currently available AD biomarkers are poor surrogate markers of αSyn pathology. Consequently, αSAA provides complementary biological information that cannot be inferred from conventional AD biomarker profiles. This observation has important implications for clinical practice, as it supports the addition of αSAA rather than reliance on existing AD biomarkers when αSyn co-pathology is clinically suspected.

Our study has certain limitations linked notably to its routine clinical setting. Some clinical data were unavailable, including REM sleep behaviour disorder, cardinal signs of parkinsonism, visual hallucinations, and cognitive fluctuations, restricting detailed phenotypic analyses according to αSAA status. We also lacked longitudinal follow-up, limiting the assessment of disease progression. However, this study represents one of the first large-scale evaluations of αSAA implementation in a real-world memory clinic cohort and demonstrates that high diagnostic performance can be achieved under routine clinical conditions.

In conclusion, αSAA can be successfully implemented in routine clinical practice while maintaining excellent diagnostic performance for LBD detection. Importantly, current AD CSF and blood biomarkers fail to identify patients harboring concomitant αSyn pathology, highlighting the unique and complementary biological information provided by αSAA. This capability could improve biological stratification of patients with cognitive disorders, optimize enrolment into future disease-specific therapeutic trials, and contribute to precision medicine approaches targeting α-synucleinopathies. Although emerging biomarkers such as CSF dopa decarboxylase (DDC) may provide additional information on synucleinopathies ^40^, αSAA currently remains the most robust and clinically validated method for detecting αSyn pathology in vivo.

## Acknowledgments

The work was funded by the Fondation Research Alzheimer (ALZAN and RTCALZ projets), AXA Mécénat Santé (INTERVAL Project) and the Fondation pour la Recherche Médicale (FRM, team Proteinopathies). None of the funding bodies had any role in study design, data collection, analysis, or interpretation, writing of the report, or in the decision to submit the paper for publication.

## Authors’ contribution

<u>Conceptualization</u> Joan Torrent, Germain Busto, Audrey Gabelle, Sylvain Lehmann

<u>Methodology</u> Marie Duchiron, Joan Torrent, Etienne Mondesert, Mehdi Morchikh, Morgan Dornadic, Constance Delaby, Christophe Hirtz, Genevieve Barnier-Figue, Karim Bennys, Sylvain Lehmann

<u>Software</u> Etienne Mondesert, Sylvain Lehmann

<u>Data curation</u> Cédric Turpinat, Germain Busto, Lou Thizy, Genevieve Barnier-Figue, Florence Perrein, Snejana Jurici, Karim Bennys, Sylvain Lehmann

<u>Investigation</u> Océane Jourdan, Marie Duchiron, Cédric Turpinat, Germain Busto, Constance Delaby, Christophe Hirtz, Genevieve Barnier-Figue, Florence Perrein, Snejana Jurici, Audrey Gabelle, Karim Bennys, Sylvain Lehmann

<u>Validation</u> Océane Jourdan, Cédric Turpinat, Etienne Mondesert, Germain Busto, Mehdi Morchikh, Lou Thizy, Sylvain Lehmann

<u>Formal analysis</u> Etienne Mondesert, Sylvain Lehmann

<u>Supervision</u> Sylvain Lehmann

<u>Funding acquisition</u> Constance Delaby, Christophe Hirtz, Audrey Gabelle, Sylvain Lehmann

<u>Visualization</u> Océane Jourdan, Mehdi Morchikh, Sylvain Lehmann

Project administration Sylvain Lehmann

<u>Resources</u> Sylvain Lehmann

<u>Writing - original draft</u> Océane Jourdan, Sylvain Lehmann

<u>Writing - review & editing</u> Océane Jourdan, Marie Duchiron, Joan Torrent, Cédric Turpinat, Etienne Mondesert, Germain Busto, Mehdi Morchikh, Morgan Dornadic, Constance Delaby, Christophe Hirtz, Lou Thizy, Genevieve Barnier-Figue, Florence Perrein, Snejana Jurici, Audrey Gabelle, Karim Bennys, Sylvain Lehmann

## Data Availability

Data and informed consent forms are available upon request (CHU Montpellier). Requests will be considered by each study investigator, based on the information provided by the requester, regarding the study and analysis plan. If the use is appropriate, a data sharing agreement will be put in place before distributing a fully de-identified version of the dataset, including the data dictionary used for analysis with individual participant data.

## Ethical approval

All participants gave written informed consent to be part of the study. The ALZAN study was approved by the “Comité de Protection des Personnes Nord Ouest IV” under # 2022-A00565-38. Clinical trial registration: NCT05427448.

