## Supplementary material for "Routine implementation of α-synuclein Seed Amplification Assays reveals high diagnostic performance and the limited value of Alzheimer disease fluid biomarkers for detecting α-synuclein co-pathology"

### Supplementary Table I: Diagnostic composition of the "Other" clinical group (n=138).

Breakdown of clinical diagnoses included in the "Other" diagnostic group (n=138) of the ALZAN cohort.

| Category | Diagnoses | n |
| --- | --- | --- |
| Atypical tauopathies | PSP, CBS, PPA | 14 |
| Vascular | Vascular dementia, CAA | 19 |
| Psychiatric disorder | Depression, bipolar disorder, ADHD, anxiety disorders | 26 |
| SCI | Subjective cognitive impairment | 11 |
| MCI | Mild cognitive impairment | 21 |
| Inflammatory/infectious | Autoimmune encephalitis, herpetic encephalitis, limbic encephalitis, CIDP | 5 |
| No cognitive impairment | / | 4 |
| Atypical parkinsonian syndromes | / | 3 |
| Non-neurodegenerative causes | NPH, epilepsy, headache, dural fistula, vertigo, OSAS, alcohol-related, lobar hematoma | 26 |
| Other neurodegenerative causes | CJD, cerebellar syndrome, LATE, Korsakoff syndrome | 9 |

**Abbreviation**: PSP, Progressive Supranuclear Palsy, CBS, Corticobasal Syndrome, PPA, Primary Progressive Aphasia, CAA, Cerebral Amyloid Angiopathy, ADHD, Attention Deficit Hyperactivity Disorder, SCI, Subjective cognitive impairment, MCI, Mild cognitive impairment, CIDP, Chronic Inflammatory Demyelinating polyradiculoneuropathy, NPH, Normal Pressure Hydrocephalus, OSAS, Obstructive Sleep Apnea Syndrome, CJD, Creutzfeldt-Jakob Disease, LATE, Limbic-predominant Age-related TDP-43 Encephalopathy

### Supplementary Figure 1: αSAA repeatability and reproducibility

**αSAA repeatability**

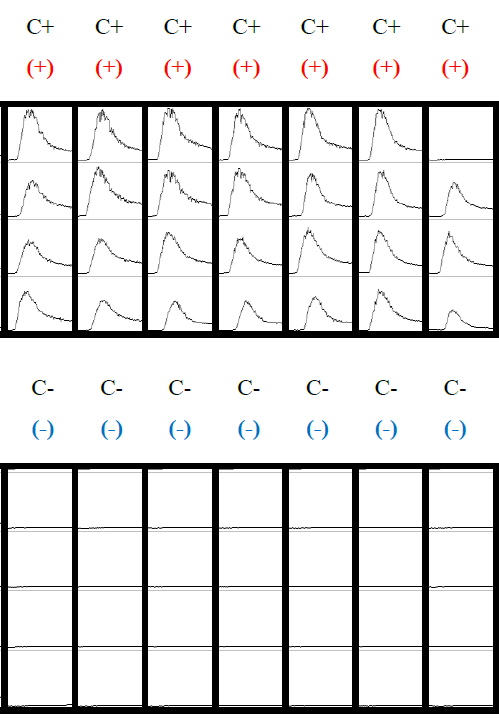
Repeatability was evaluated by analyzing positive and negative controls deposited seven times on the same plate during the same day, each in four replicates, under identical experimental conditions. As illustrated in the figure below, all positive controls showed at least 3 amplifications out of 4 replicates. Consequently, all eight repetitions of the positive control were considered positive, corresponding to a repeatability of 100% (n = 7/7). When considering each well individually, 27 out of 28 wells were positive, corresponding to a repeatability of 96.4%. Similarly, no amplification was observed in the seven additional replicates of the negative control, indicating a repeatability of 100%, both at the sample level and at the well level.

**Abbreviation:** C, Control

**αSAA reproducibility**

Reproducibility was evaluated across more than 30 independent assays. For each assay, positive and negative controls were each tested in four replicates. Reproducibility was therefore assessed in 35 assays for the positive control and in 31 assays for the negative control. For the negative control, reproducibility was 93.6%, with 29 negative results out of 31 assays. For the positive control, reproducibility reached 97.1% (n = 34/35), with 34 positive results out of 35 assays.

**αSAA inter-operator variability**

Four operators were tested. The operator 1, who performed the largest number of αSAA (n = 58) exhibited slightly higher variability (93%) compared with 100% reproducibility for the other operators. However, inter-operator variability remained satisfactory.

|  | Operator 1 (n=58) | Operator 2 (n=4) | Operator 3 (n=2) | Operator 4 (n=3) |
| --- | --- | --- | --- | --- |
| Positive control | 93.2% | 100% | 100% | 100% |
| Negative control | 93% | 100% | 100% | 100% |

### Supplementary Table 2: αSAA method comparison

This table directly compares the αSAA results obtained simultaneously in the laboratories of Lyon and Montpellier. For each sample, αSAA results are presented as the number of positive replicates over the total number of replicates tested. A binary qualitative interpretation (P for positive, N for negative) is provided for each laboratory. The concordance column indicates agreement (Y) or disagreement (N) between the qualitative results from the two sites. Overall qualitative concordance between the Lyon and Montpellier laboratories was high, with 45 concordant results out of 48 samples, corresponding to an overall concordance rate of 93.8%.

| Id | Initial Diagnosis | Lyon | Montpellier | Lyon | Montpellier | Concordance |
| --- | --- | --- | --- | --- | --- | --- |
| LYO3 | AD | 0/4 | 1/8 | N | N | Y |
| LYO8 | IP | 4/4 | 3/4 | P | P | Y |
| LYO14 | AD / LBD | 0/4 | 0/4 | N | N | Y |
| LYO2 | AD | 1/8 | 0/4 | N | N | Y |
| LYO4 | LBD | 4/4 | 4/4 | P | P | Y |
| LYO10 | IP | 3/4 | 3/4 | P | P | Y |
| LYO7 | LBD | 1/8 | 0/4 | N | N | Y |
| LYO1 | AD | 3/8 | 0/4 | P | N | Y |
| LYO13 | AD / LBD | 4/4 | 2/4 | P | P | Y |
| LYO9 | MPI | 0/4 | 3/4 | N | P | N |
| LYO12 | DFT | 0/4 | 1/8 | N | N | Y |
| 14300 | MCI | 1/8 | 0/4 | N | N | Y |
| 14398 | MCI | 0/4 | 0/4 | N | N | Y |
| 14454 | AD | 4/4 | 4/4 | P | P | Y |
| 14832 | AD | 0/4 | 1/8 | N | N | Y |
| 15589 | NPH | 0/4 | 0/4 | N | N | Y |
| 15717 | AD / LBD | 2/4 | 2/4 | P | P | Y |
| 16057 | MCI | 2/4 | 2/4 | P | P | Y |
| 16384 | Parkinsonism | 2/4 | 3/4 | P | P | Y |
| 16442 | MCI | 4/4 | 4/4 | P | P | Y |
| 70676 | MCI | 0/4 | 0/4 | N | N | Y |
| 102417 | AD / LBD | 4/4 | 3/4 | P | P | Y |
| 402753 | AD / LBD | 4/4 | 3/4 | P | P | Y |
| 102389 | LBD | 4/4 | 4/4 | P | P | Y |
| 403110 | AD / LBD | 3/8 | 2/4 | P | P | Y |
| 403199 | MCI | 1/8 | 0/4 | N | N | Y |
| 202074 | Sarcoidosis | 0/4 | 1/8 | N | N | Y |
| 301964 | LBD | 0/4 | 1/8 | N | N | Y |
| 501902 | MCI | 0/4 | 0/4 | N | N | Y |
| 201791 | AD | 0/4 | 0/4 | N | N | Y |
| 302041 | LBD | 0/4 | 2/4 | N | P | N |
| 402029 | LBD | 0/4 | 0/4 | N | N | Y |
| 202213 | LBD | 3/4 | 4/4 | P | P | Y |
| 202143 | LBD | 3/8 | 2/4 | P | P | Y |
| 6918 | DCL | 4/4 | 4/4 | P | P | Y |
| 7609 | APP / LBD | 4/4 | 4/4 | P | P | Y |
| 8268 | AD / LBD | 3/4 | 4/4 | P | P | Y |
| 9154 | FTD / LBD | 4/4 | 4/4 | P | P | Y |
| 491 | AD | 0/4 | 0/4 | N | N | Y |
| 468 | AD | 0/4 | 0/4 | N | N | Y |
| 2585 | AD | 0/4 | 0/4 | N | N | Y |
| 3634 | AD | 4/4 | 3/4 | P | P | Y |
| 1103L211250 | LBD | 0/4 | 0/4 | N | N | Y |
| 1109L070991 | LBD | 2/4 | 3/4 | P | P | Y |
| 1109L080878 | LBD | 4/4 | 4/4 | P | P | Y |
| 1203L211228 | LBD | 0/4 | 0/4 | N | N | Y |
| 1201L301174 | LBD / vasc. | 0/4 | 2/4 | N | P | N |
| 1004L211098 | LBD | 0/4 | 0/4 | N | N | Y |

**Abbreviation:** AD, Alzheimer’s disease; LBD, Lewy body disease; DCL, dementia with Lewy bodies; IP, idiopathic Parkinson’s disease; MCI, mild cognitive impairment; MPI, multiple parkinsonian syndromes; FTD, frontotemporal dementia; NPH, normal pressure hydrocephalus; vasc., vascular; P, positive; N, negative; Y, yes; N (Concordance), no.

### Supplementary Figure 2: ROC curve

ROC curves were generated for several plasma biomarkers to evaluate their ability to detect AD. Supplementary Figure 1 presents the individual diagnostic performance of pTau217, Aβ42/40 ratio, GFAP, and NfL. The biomarker pTau217 showed the best diagnostic performance, with an AUC of 0.902 (95% CI: 0.871–0.933), followed by Aβ42/40 ratio (AUC = 0.803; 95% CI: 0.758–0.848). GFAP showed intermediate performance (AUC = 0.711; 95% CI: 0.659–0.762), whereas NfL demonstrated limited discriminative ability, with an AUC of 0.549 (95% CI: 0.489–0.608).

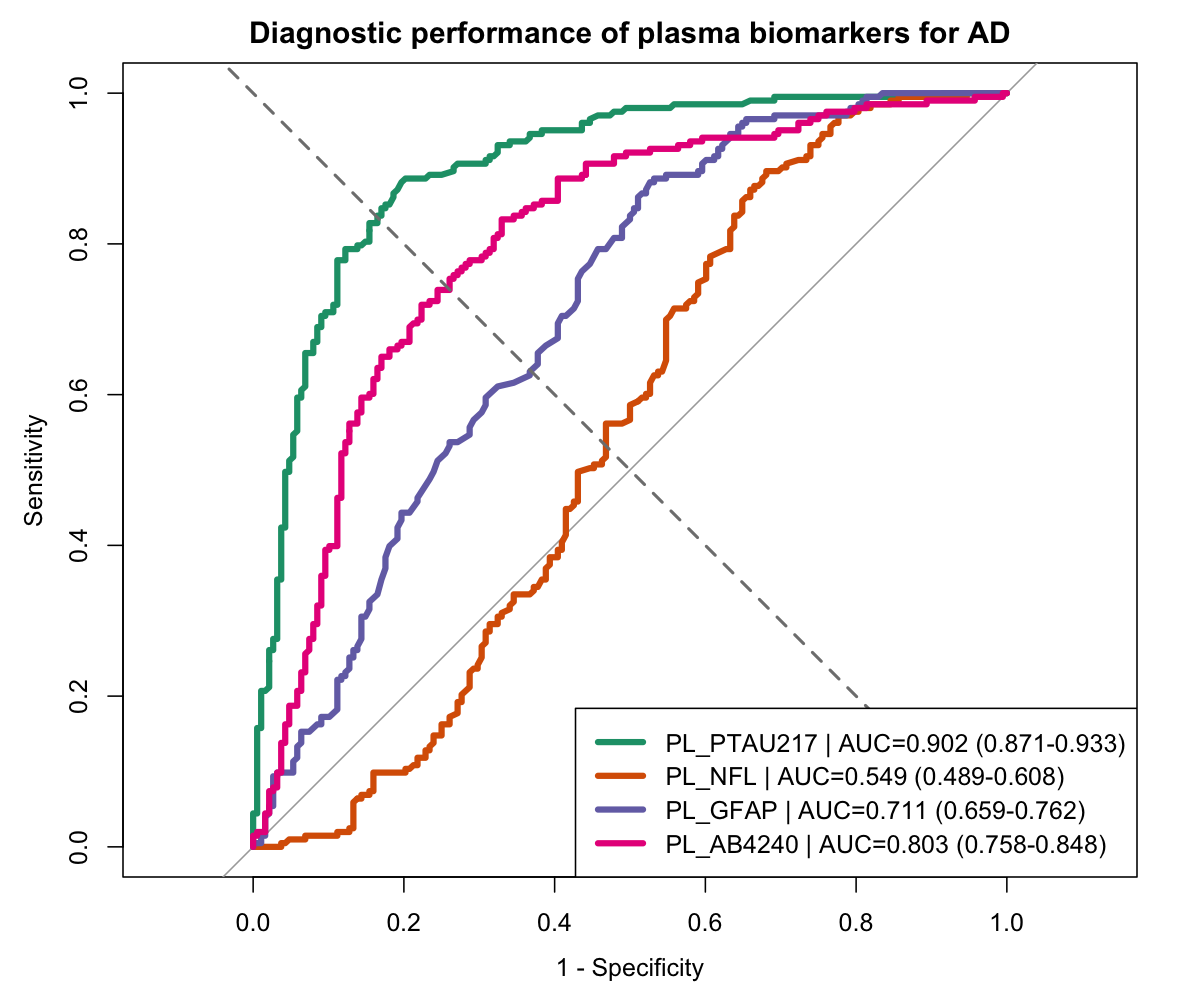

**Abbreviation:** AD, Alzheimer’s disease; Pl, Plasma; ptau, phosphorylated tau; NfL, neurofilament light chain; GFAP, glial fibrillary acidic protein.; Aβ42/40, amyloid β-peptide ratio; AUC, Area Under the Curve

### Supplementary Table 3: Demographic, cognitive, and biological biomarker results between αSAA- and αSAA+ patients (n = 398).

Data are presented as mean (M ± SD) for continuous variables and n (%) for categorical variables. Differences between αSAA− and αSAA+ patients were assessed using Student’s-t-test for continuous variables and χ² test for categorical variables. P-values are shown in the last column (*p < 0.05; **p < 0.01).

| Criteria | αSAA- (n=335) | αSAA+ (n=63) | p-value |
| --- | --- | --- | --- |
| Age (years), M (SD) | 70.5 (10.9) | 73.9 (6.8) | *p*=0.0013 ** |
| Sexe, female, n (%) | 183 (54.6) | 26 (41.3) | *p*=0.0514 |
| BMI (kg/m2), M (SD) | 24.8 (4.1) | 23.9 (3.8) | *p*=0.1394 |
| MMSE (/30), M (SD) | 23.4 (4.6) | 21.6 (4.3) | *p*=0.0085 ** |
| eGFR (mL/min/1.73m^2^), M (SD) | 84.9 (16.2) | 83.7 (13.5) | *p*=0.5502 |
| CSF Aβ40 (pg/mL), M (SD) | 10777 (3822) | 11273 (4277) | *p*=0.3938 |
| CSF Aβ42 (pg/mL), M (SD) | 748.7 (366.1) | 650.2 (291.2) | *p*=0.0203 * |
| CSF Aβ42/40 (%),M (SD) | 7.3 (2.8) | 6.2 (2.6) | *p*=0.0051 ** |
| CSF tau (pg/mL), M (SD) | 483.3 (358.8) | 517.6 (320.5) | *p*=0.4468 |
| CSF ptau181 (pg/mL), M (SD) | 69.9 (58.7) | 78.2 (54.5) | *p*=0.2752 |
| Pl GFAP (pg/mL), M (SD) | 132.7 (65.2) | 148.7 (53.8) | *p*=0.0395 * |
| Pl ptau217 (pg/mL), M (SD) | 0.41 (0.43) | 0.39 (0.35) | *p*=0.6697 |
| Pl Aβ40 (pg/mL), M (SD) | 296.6 (53.7) | 298.3 (47.4) | *p*=0.7986 |
| Pl Aβ42 (pg/mL), M (SD) | 34.9 (8.7) | 35.1 (6.9) | *p*=0.9515 |
| Pl Aβ42/40 (%),M (SD) | 11.9 (2.4) | 12.1 (4.4) | *p*=0.6507 |
| Pl ptau181 (pg/mL), M (SD) | 1.3 (0.77) | 1.3 (0.64) | *p*=0.9181 |
| Pl Nfl (pg/mL), M (SD) | 4.5 (5.2) | 4.1 (2.1) | *p*=0.2638 |
| APOE ε4 carrier, n (%) | 140 (41.8) | 31 (49.2) | *p*=0.2754 |

**Abbreviation**: AD, Alzheimer's disease; LBD, Lewy body dementia; FTD, frontotemporal dementia; BMI, body mass index; MMSE, Mini-Mental State Examination; eGFR, estimated glomerular filtration rate; CSF, cerebrospinal fluid; Pl, plasma; ptau, phosphorylated tau; NfL, neurofilament light chain; αSAA, α-synuclein seed amplification assay.
